# Is nasopharyngeal swab comparable with nasopharyngeal aspirate to detect SARS-CoV-2 in children?

**DOI:** 10.1101/2020.07.02.20142521

**Authors:** Ester Capecchi, Giada Maria Di Pietro, Ester Luconi

## Abstract

The tests currently used for the direct identification of SARS-CoV-2 include specimens taken from the upper and the lower respiratory tract.

In our paediatric department all children undergo both nasopharyngeal swab and nasopharyngeal aspirate, performed from both nostrils, on admission and after 24 hours.

We decided to compare these two methods of detection of SARS-CoV-2. Considering nasopharyngeal aspirate as the gold standard, we calculated sensitivities and specificities of nasopharyngeal swab.

Based on our results, we suggest to prefer the collection of aspirates whenever possible.

## To the Editor

In December 2019 appeared in China a novel coronavirus, designated as SARS-CoV-2, responsible for a pandemic respiratory disease, known as coronavirus Disease (COVID-19), with the Italian outbreak from February 2020. Children appear to have milder symptoms and less severe disease^1^. The tests currently used for the direct identification of SARS-CoV-2 include specimens taken from the upper and the lower respiratory tract^2,3^. Since the use of nasopharyngeal aspirate (NPA) seemed to be better than nasopharyngeal swab (NS) to identify respiratory virus in paediatrics^4,5^ we decided to compare these methods in detecting SARS-CoV-2 in children.

Children hospitalized in our paediatric department underwent NS (Copan-503CS01 nasopharingeal flocked swab) and NPA (Medicoplast mucus extractor 440-ch08), performed from both nostrils, on admission and after 24 hours. SARS-CoV-2 RNA was extracted from the paired samples through nucleic acid amplification, using the RT-PCR.

From March 13th to May 22nd, 300 paired specimens (NS/NPA) collected from 136 patients (134 hospitalized and 2 outpatients) were tested for SARS-CoV-2.

For clinical aims, we considered positive, to SARS-CoV-2 every patient whose NPA or NS or NPA/NS resulted positive or weak positive.

Out of the 134 patients hospitalized, 18 children tested positive (prevalence 13.4%, 95% CI: 8.2%-20.4%); among the latter, 13 of them and 2 outpatient children were followed collecting their paired specimens until both resulted negative 24 hours apart.

Of the 300 paired specimens evaluated: 276 were concordant, 24 were discordant, so the naïve concordance was 92.0% (95% CI: 88.3%-94.6%) with Cohen’s kappa (K) 0.63. Among the paired specimens whose NPA resulted positive, 41.9% (95% CI: 28.2%-56.9%) had NS negative; while among the paired specimens whose NPA resulted negative, 2.3% (95% CI: 1.1%-5.1%) had NS positive.

Considering NPA as the gold standard for detection of SARS-CoV-2, we calculated sensitivities and specificities of NS. The overall sensitivity of NS was 58.1% (95% CI: 43.1%-71.8%) and the specificity was 97.7% (95% CI: 94.9-98.9%). Since the different practice in specimen collection, we divided our cohort according to the children’s age (<6 or ≥ 6 years, Table 1). Among children under 6 years, the concordance was K=0.67. Regarding children of 6 years or older, the concordance was K=0.60.

**Table 1.**
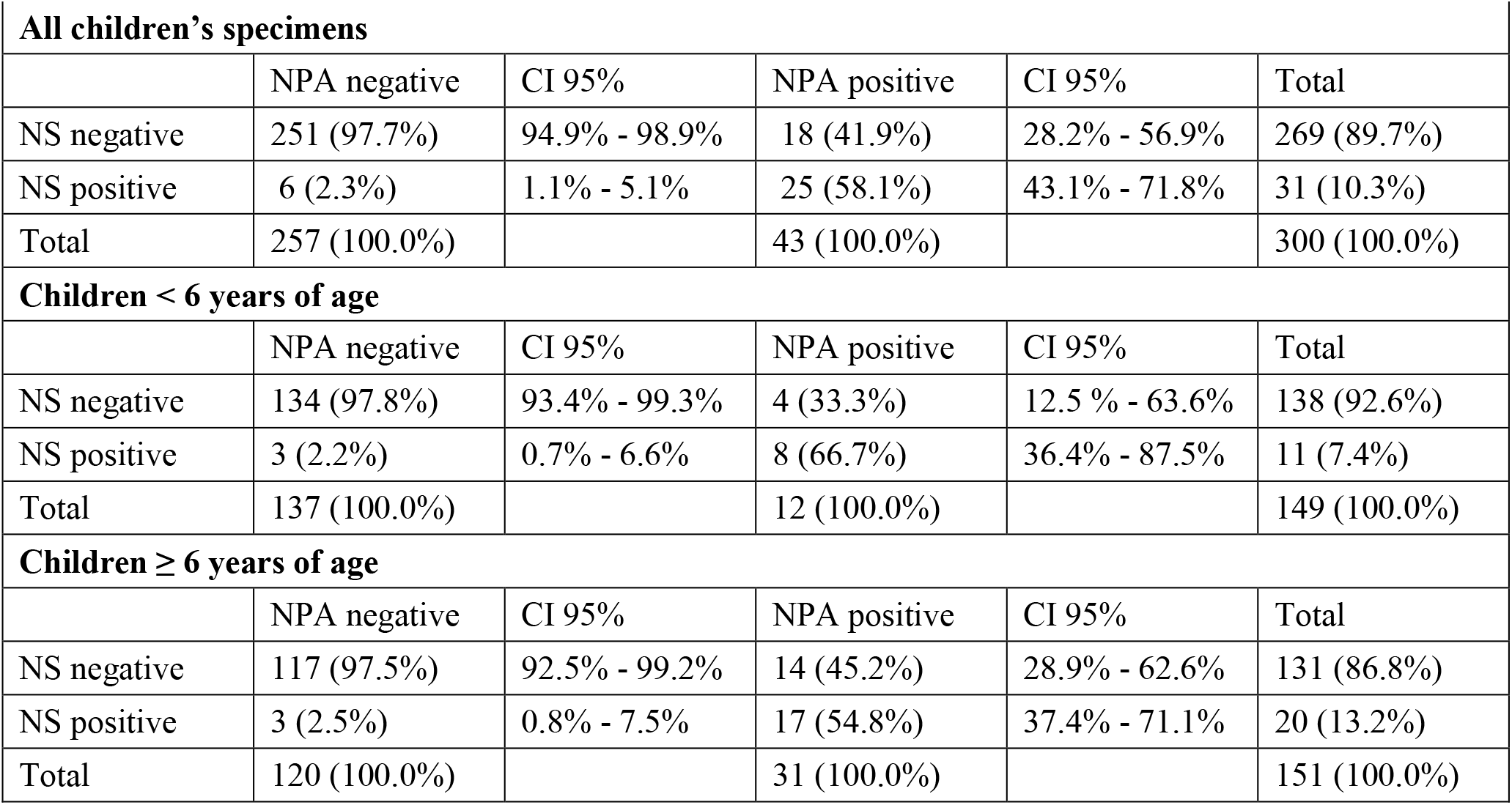
NS and NPA results.

| **Table 1. NS and NPA results** |  |  |  |  |  |
| --- | --- | --- | --- | --- | --- |
| **All children's specimens** |  |  |  |  |  |
|  | NPA negative | CI 95% | NPA positive | CI 95% | Total |
| NS negative | 251 (97.7%) | 94.9% - 98.9% | 18 (41.9%) | 28.2% - 56.9% | 269 (89.7%) |
| NS positive | 6 (2.3%) | 1.1% - 5.1% | 25 (58.1%) | 43.1% - 71.8% | 31 (10.3%) |
| Total | 257 (100.0%) |  | 43 (100.0%) |  | 300 (100.0%) |
| **Children < 6 years of age** |  |  |  |  |  |
|  | NPA negative | CI 95% | NPA positive | CI 95% | Total |
| NS negative | 134 (97.8%) | 93.4% - 99.3% | 4 (33.3%) | 12.5 % - 63.6% | 138 (92.6%) |
| NS positive | 3 (2.2%) | 0.7% - 6.6% | 8 (66.7%) | 36.4% - 87.5% | 11 (7.4%) |
| Total | 137 (100.0%) |  | 12 (100.0%) |  | 149 (100.0%) |
| **Children ≥ 6 years of age** |  |  |  |  |  |
|  | NPA negative | CI 95% | NPA positive | CI 95% | Total |
| NS negative | 117 (97.5%) | 92.5% - 99.2% | 14 (45.2%) | 28.9% - 62.6% | 131 (86.8%) |
| NS positive | 3 (2.5%) | 0.8% - 7.5% | 17 (54.8%) | 37.4% - 71.1% | 20 (13.2%) |
| Total | 120 (100.0%) |  | 31 (100.0%) |  | 151 (100.0%) |

The NS has in any case a low sensitivity in detecting SARS-CoV-2 in children when referred to NPA. Our results, the first we know are available, suggest to prefer the collection of NPA whenever possible for the detection of SARS-CoV-2 in children.

## Data Availability

all data of the study are available

